# Diversity of *CFTR* variants across ancestries characterized using 454,727 UK biobank whole exome sequences

**DOI:** 10.1101/2023.03.10.23287070

**Authors:** Justin E. Ideozu, Mengzhen Liu, Bridget M. Riley-Gillis, Sri R. Paladugu, Fedik Rahimov, Preethi Krishnan, Rakesh Tripathi, Patrick Dorr, Hara Levy, Ashvani Singh, Jeffery F. Waring, Aparna Vasanthakumar

## Abstract

Limited understanding of the diversity of *CFTR* variants across ancestries hampers efforts to advance molecular diagnosis of cystic fibrosis (CF). The consequences pose a risk of delayed diagnoses and subsequently worsened health outcomes for patients. Characterizing the distribution of *CFTR* variants across ancestries is, therefore, critical for revolutionizing molecular diagnoses of CF. By interrogating 454,727 UK biobank whole exome Sequences, we detected over 4,000 *CFTR* variants, including novel ancestry-specific variants, across six ancestries. F508del was the most prevalent CF-causing variant found across the ancestries, except in East Asia, where V520F was the most prevalent. Sixteen participants had two CF-causing variants, with two diagnosed as CF. We found 154 participants with a CF-causing and varying clinical consequences (VCC) variant. Overall, participants with multiple clinically relevant variants reported indications significantly associated with CF and its pulmonary phenotypes [Bonferroni adjusted p<0.05]. Potentially, participants with classical CF phenotypes may benefit from current CF therapies.

## Introduction

In cystic fibrosis (CF), pathogenic variants in the cystic fibrosis transmembrane conductance regulator (*CFTR*) gene results in system-wide debilitating consequences^1^. Current CFTR modulator therapies targeting the basic CFTR defects have yielded both short- and long-term clinical benefits, but they are approved only for patients with certain *CFTR* variants^2,3^. Even within these patients, the response to current CFTR modulator therapies can be variable. Recent advances in genetic technologies have led to the discovery of many CF-causing variants among populations of European ancestry. Although these findings have led to a better understanding of the disease prevalence, and improved diagnoses^4^, there is limited knowledge about the distribution of ancestry-specific *CFTR* variants. Characterizing the broad spectrum of pathogenic and non-pathogenic *CFTR* variants across ancestries holds promise to revolutionize molecular diagnoses of CF and could enable wider access to personalized CFTR modulator therapies.

Most studies that have surveyed the diversity of *CFTR* variants have largely focused on studying persons with CF from European populations^5,6^. This is perhaps driven by the fact that CF predominantly affects individuals from European ancestry, and the disease is less frequent in people from other ancestries. Survey reports from a diverse US population shows CF affects individuals of European ancestry (1:3000 live birth) relative to people from other ethnicities, including Africans (1:15000), Asians (1:35,000), and Native Americans (1:10,900)^7,8^. Globally, CF is presumed to affect about 100,000 people and F508del accounts for most of the cases^6,9^. Recent reports suggest the spectrum of CF-causing variants vary across ancestries and countries^7,10^. CF-causing variants commonly reported in other ethnic groups, include 3120+1G>A (Africans)^10^, p.W1282X (Ashkenazim)^6,11^, and p.G970D (Chinese)^12^. Although these studies have shed insights on common CF-causing variants in non-European populations, carrier screening panels do not often capture the dominant CF-causing variants in some ethnicities^13^. Thus, the prevalence of the disease in other ethnicities, and worldwide, are possibly underestimated.

Diagnosis of CF can be based on the results of a patients’ pilocarpine iontophoresis sweat test (sweat chloride ≥ 60 mmol/L) and/or molecular genetic testing (positive for up to two CF-causing variants)^14^. One of the advantages of molecular genetic testing is that it could detect both symptomatic and pre-symptomatic individuals, including the potential to determine a patients’ eligibility for current CFTR modulator therapies. A major drawback of solo reliance on symptomatic diagnoses is that often patients are diagnosed when they are already presenting symptoms, and delayed diagnosis negatively impacts the patients’ health outcomes15. Early diagnosis and interventions, particularly with triple CFTR therapy, results in improved clinical outcomes and could profoundly affect the trajectory of CF lung disease^16^. Carriers of rare pathogenic variants not included in carrier screening panels are more likely to have delayed diagnosis which poses a risk of increased morbidity. Thus, it has become critical to characterize the broad spectrum of pathogenic *CFTR* variants across ancestral populations.

Determining the clinical and functional relevance of *CFTR* variants is such an inundated task that requires critical examination of the variants’ impact on an individual’s phenotype. This is compounded by the clinical heterogeneity often seen in patients with the same *CFTR* genotype. Efforts driven by CFTR1/2 have led to the identification of 2,110 *CFTR* variants [http://www.genet.sickkids.on.ca]. Of these, 401 are CF-causing while 49 are variants of varying clinical consequences (VCC), according to the latest CFTR2 annotation [https://cftr2.org]. The CFTR1/2 databases have led to an increased understanding of CF diagnoses and prevalence across populations. However, the variants captured from the CFTR1/2 efforts do not necessarily represent the entirety of potential variants that could possibly exist within the *CFTR* gene across ancestries. Recent annotations from TopMed Whole Genome Sequencing (WGS) efforts that includes diverse populations indicate that the *CFTR* gene is about 250kb with 56,488 variants^17^, which is longer than the often referenced 189kb length^18^. Although the vast majority of these newly detected *CFTR* variants are intronic [96%], and their clinical relevance unknown, this underscores the continued need to gain a comprehensive understanding of all pathogenic and non-pathogenic variants that possibly exists within the *CFTR* gene, including those from various ancestries.

The complexities associated with characterizing the clinical relevance of *CFTR* variants can be subdued by leveraging population databases, such as UK Biobank (UKBB), that hosts both genomics and phenotypic information of individuals. In this study, we characterized the diversity of *CFTR* variants across six ancestral populations included in the UKBB. We segregated ancestry-specific *CFTR* variants, including those that are CF-causing from uncharacterized variants. We estimated the age of certain detected CF-causing variants. For participants with two CF-causing variants or a CF-causing variant and a VCC, we performed phenotype analysis to determine the possible impact of pathogenic CF-variants on their health outcomes. Our work provides the foundation for future studies to explore the clinical relevance of the newly detected *CFTR* variants across the described ancestries.

## Methods

### Study population and whole exome sequences

The dataset used for this study is available in the UKBB public repository and was accessed under application 26041. A total of 454,787 UKBB whole exome sequences were interrogated to characterize the diversity of *CFTR* variants across ancestries. The demographics and characteristics of the UKBB participants has been described elsewhere^19^, but relevant information such as gender, age and spirometry measures were recorded for each participant. Details on the calling and quality control can be found in previous publication^19^. To maximize variant discovery, no further filters were applied except the removal of individuals that subsequently withdrew consent (final N = 454,727). Curated disease phenotypes, defined based on participants primary care and hospital in-patient (HESIN) records, were utilized for association tests with pathogenic *CFTR* genotypes.

### Ancestry designation

For ancestry designation, we opted to use the classification from the PanUKBB working group (https://pan-dev.ukbb.broadinstitute.org/) which derives the ancestry classification from two large reference datasets, 1000 Genomes Project and Human Genome Diversity Project. Using this classification, the participants were grouped into six major groups: African (AFR), American/American Admixed (AMR), Central South Asia (CSA), East Asian (EAS), European (EUR), and Middle East (MID). Individuals that did not fall neatly into the large continental classifications were classified as MIX. While this classification limits the full extent of genomic diversity in the UKBB, it permitted an estimation of variant frequencies to other populations. Individuals assigned to the MIX group were included in all other counts and analysis.

### Variant annotation

Clinically relevant variants (CF-causing and VCCs) detected across the ancestries were classified using CFTR2 annotation. CF-causing variants without an assigned Reference SNP cluster ID (rsID) were excluded since they comprised mostly of large indels and structural variants, which are often challenging to be called by variant calling tools. Functionally relevant variants (high impact variants) were annotated with Variant Effect Predictor^20^ and SnpEFF^21^ and those high impact variants not known to cause CF were recorded. The canonical transcript of *CFTR* (ENST00000003084) was used as reference for both tools. CF is an autosomal recessive disease and molecular diagnosis could be based on the possession of two or more clinically relevant variants. Thus, participants with two CF-causing variants, a CF-causing variant and a VCC, and a CF-Causing variant and a non-CF-causing high impact variant, were characterized and prioritized for further association test. Participants with two CF-causing variants were then assigned likely disease severity status (pancreatic sufficient or insufficient) based on their *CFTR* genotype as previously described^22,23.^

### Statistical association analysis

We categorized participants with certain clinical or functionally relevant variant combinations into three groups: two CF-causing variants, a CF-causing variant and a VCC, a CF-causing variant and a variant of high impact. We tested each of the groups for association with disease phenotypes using Fisher-Exact tests. As pulmonary measures such as forced expiratory volume (FEV) and forced vital capacity (FVC) could reflect pulmonary phenotype, we tested for differences in these measures between participants with two CF-causing variants and the general population. Only associations meeting a Bonferroni corrected adjusted p-value < 5% were considered as statistically significant.

### Genealogical estimation of CF-causing variants age

To estimate the age of CF-causing variants, we used data from the Atlas of Variant Age (AVA) (https://human.genome.dating/). Models using data from the 1000 Genomes Project (TGP) and the Simons Genome Diversity Project (SGDP), implemented in the Genealogical Estimation of Variant Age (GEVA) tool^24^, were deployed. For variant age estimations, joint mutational and recombinational clocks were considered using an average generation time of 25 years. Estimated age of variants within the *CFTR* gene locus (Chr7: 117,095,837 – 117,366,025) were then reported.

## Result

### Over 4000 *CFTR* variants identified across ancestries

We interrogated 454,787 whole exome sequences available in the UKBB to characterize the diversity of *CFTR* variants across six ancestries [AFR, CSA, EAS, EUR, AMR, and MID] and an uncharacterized group (MIX). The median age across all ancestries was greater than 60 years. Except for CSA and MID where over half of the characterized participants were males, most of the ancestries were dominated by females (Table 1). Overall, we detected 4,193 variants across all ancestries in the UKBB, with V470M emerging as the most common variant (Table S1). The *CFTR* variants represented a diverse range of variant types, but over 50% were intronic (Fig. 1) The highest number of *CFTR* variants were detected in Europeans [n=3,192] while the AMR group had the least number of *CFTR* variants [n=151]. Across the remaining ancestries [CSA, AFR, EAS, and MID], the number of *CFTR* variants detected [471, 417, 266, and 222 respectively] were also much lower when compared to those detected in Europeans (Table 1). Many of the variants detected in this study have never been reported as variants in *CFTR*. Next, we explored if there were *CFTR* variants uniquely represented across the ancestries.

**Table 1.** Diversity of CFTR variants across ancestries

| Approximate global ancestry | Sample Size, n (%) | Gender: Male, n (%) | Age in Years, Median (IQR) | CFTR Variants, n (%) | Unique CFTR Variants, n (%) |
| --- | --- | --- | --- | --- | --- |
| African (AFR) | 6278 (1.4%) | 2582 (41%) | 62 (57, 70) | 417 (9.9%) | 138 (33.1%) |
| Central South Asia (CSA) | 8470 (1.9%) | 4572 (54%) | 65 (58, 72) | 471 (11.2%) | 177 (37.6%) |
| East Asia (EAS) | 2608 (0.6%) | 886 (34%) | 63 (57, 70) | 266 (6.3%) | 104 (39.1%) |
| European (EUR) | 397,073 (87.3%) | 182164 (46%) | 70 (62, 75) | 3192 (76.1%) | 2212 (69.3%) |
| American/Admixed American (AMR) | 950 (0.2%) | 334 (35%) | 63 (57, 71) | 151 (3.6%) | 23 (15.2%) |
| Middle East (MID) | 1498 (0.3%) | 872 (58%) | 63 (57, 70) | 222 (5.3%) | 46 (20.7%) |
| MIX | 37850 (8.3%) | 16450 (43%) | 67 (60, 67) | 1333 (31.8%) | 381 (28.6%) |

**Fig. 1:**
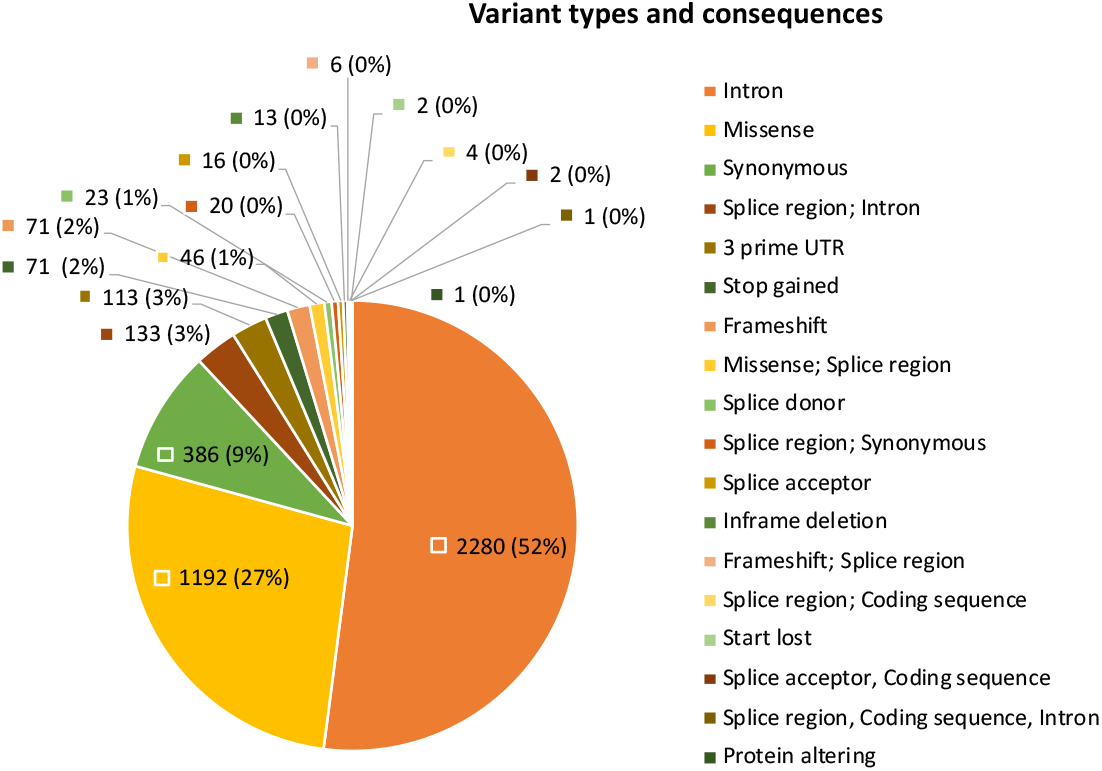
Most *CFTR* variants are intronic. Several variant types with varying consequences were identified in the *CFTR* gene. The labels in the chart represent the number and percentage of the Intronic variants (highlighted in orange) clearly dominated over half of the captured variants despite Whole Exome Sequencing technology capturing mostly exomes.

Interestingly, we found several *CFTR* variants that tend to be ancestry specific (Fig. 2, Table S2). The highest number of unique *CFTR* variants [2212/3192] was found in Europeans, while the Hispanic/Admixed American group had the least number of unique *CFTR* variants [23/151]. About 8.3% [37850/454,727] of the overall study population did not fall under a specific ancestry and were categorized as the Mixed group (MIX). The median (interquartile range [IQR] age for the MIX population was 67 (60, 67), with males accounting for 43% of the population. Within the MIX group, we detected a total of 1333 *CFTR* variants and 28.6% of these variants [n=381] were uniquely found in their population (Table 1).

**Fig. 2:**
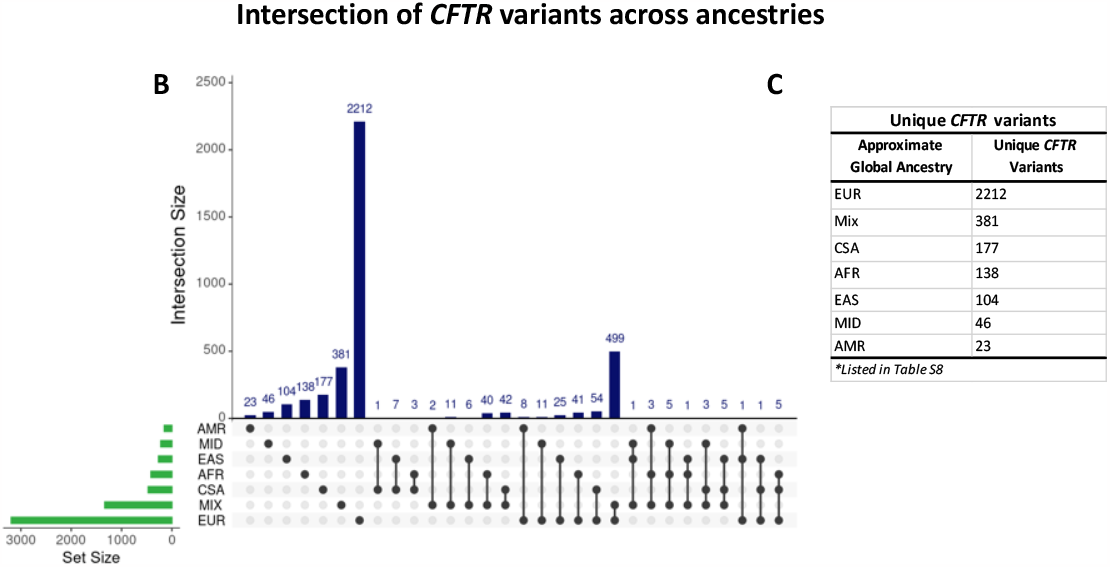
Ancestry-specific variants characterized. (A)Graph of total number of variants (x-axis; Set Size) detected in each ancestry (y-axis). **B)** Intersection of variants detected across the ancestries. Each column corresponds to the number of variants in each intersection. Ancestries present in each intersection are represented by the black dots. Intersections with single dots represents ancestry-specific variants. **C)** A list of ancestry-specific variants. The highest number of unique *CFTR* variants (2212/3192) was detected in Europeans.

### CF-causing variants across ancestries

We explored the global distribution of CF-causing variants, variants of varying clinical consequences (VCC), and high impact variants. As shown in Table 2, we found varying distribution of CF-causing (Table S3) and VCC (Table S4) variants across the ancestries. The highest number of CF-causing variants were detected in the EUR population [116/154]. These variants accounted for 75% of all CF-causing variants with RSIDs captured in the study. Nearly 50% of the CF-causing variants were found in the MIX population, but lower proportions (<15%) were reported across the other specific ancestries. The AMR group with the least number of detected *CFTR* variants [151/4193] also had the least number [7/154] of the CF-causing variants (Table 2). F508del was the most prevalent CF-causing variant observed in all ancestries, except in EAS where V520F was the most prevalent (Fig. 3). While F508del represented ∼90% of the CF-causing variants found in Europeans, our findings indicate its prevalence was not as high in other ancestries. We further explored whether there were any CF-causing variants that were potentially ancestry-specific. As shown in Figure 4, all ancestries had unique CF-causing variants, but EUR possessed the highest number [n=50] of ancestry-specific CF-causing variants while four groups (AMR, CSA, EAS, and MID) had only one ancestry-specific variant each (Table S5). We also detected VCC variants in all ancestries [Table S4]. The number of VCC variants detected were smaller than the number of CF-causing variants detected across the ancestries, except for AMR and MID populations. R74W was one of the most common variants detected across the ancestries. Except for EUR, R74W featured among the top 5 most common VCC reported in all ancestries. Other VCCs that made the common list included R117H, D1270N, and L967S (Table S6). Interestingly, we found several high impact variants that are not known to be CF-causing or clinically relevant in each ancestry. The highest number of these variants [150/200] were detected in the EUR population. Overall, high impact variants accounted for only a small fraction of the total number of variants reported in each ancestry [≤ 5%] (Table S7) and many of these [127/200] are uncharacterized in CF (Table S8).

**Table 2.** Number of participants with CF-causing variants across ancestries

| Ancestries | Sample Size in UKBB | CFTR Variants | CF-Causing | Varying Clinical Consequences (VCC) | High Impact Variants |
| --- | --- | --- | --- | --- | --- |
| AFR | 6278 | 417 | 22 | 12 | 16 |
| AMR | 950 | 151 | 7 | 10 | 4 |
| CSA | 8470 | 471 | 17 | 6 | 15 |
| EAS | 2608 | 266 | 8 | 6 | 4 |
| EUR | 397,073 | 3,192 | 116 | 29 | 150 |
| MID | 1498 | 222 | 8 | 12 | 7 |
| MIX | 37850 | 1,333 | 85 | 29 | 70 |

**Fig. 3:**
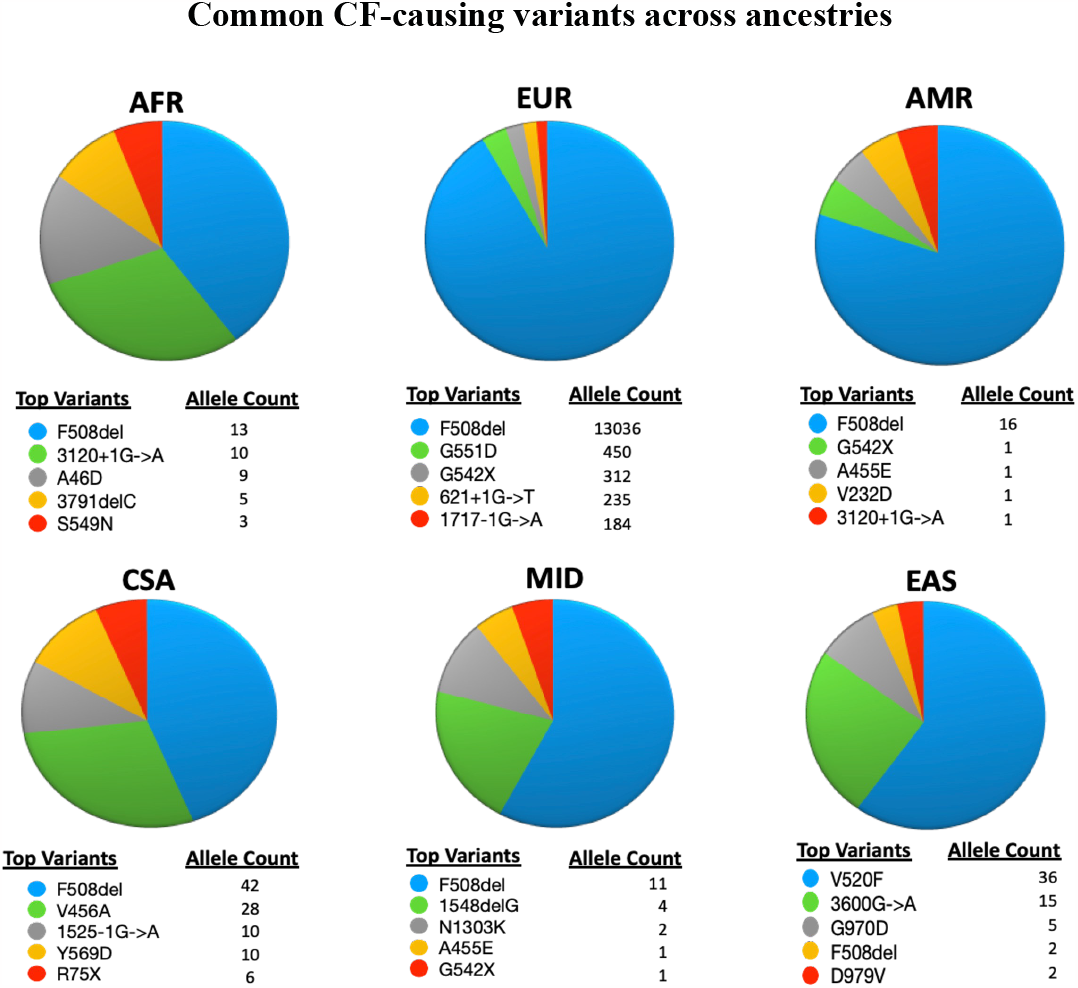
Common CF-causing variants in each ancestry represented in pie charts. For each ancestry, slices vary by color, with blue representing the most abundant CF-causing variant. F508del was the most common CF-causing variant detected in all ancestries, except for East Asia where V520F was the most common.

**Fig. 4:**
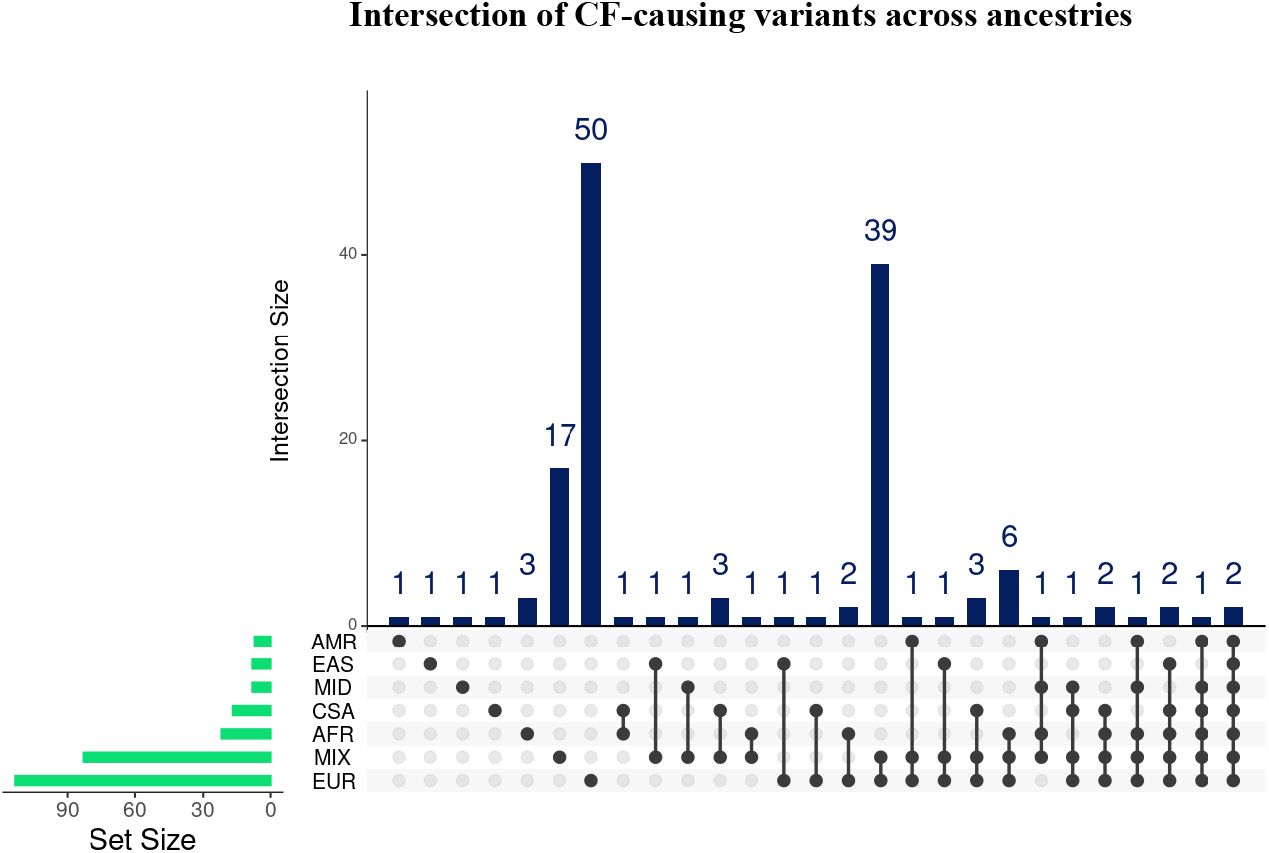
Ancestry-specific CF-causing variants characterized. (A)Graph of total number of variants (x-axis; Set Size) detected in each ancestry (y-axis). **B)** Intersection of variants detected across the ancestries. Each column corresponds to the number of variants in each intersection. Ancestries present in each intersection are represented by the black dots. Intersections with single dots represents ancestry-specific CF-causing variants. **C)** A list of ancestry-specific CF-causing variants. The highest number of unique CF-causing variants (n=50) was detected in Europeans.

### More than one CF-causing variants detected in participants

With CF being an autosomal recessive disease, we explored the prevalence of participants with two CF-causing variants and those heterozygous for a CF-causing variant and a VCC or high impact variant. We found 16 UKBB participants possessing at least two CF-causing variants (Table 3). These participants were found exclusively in the CSA [n=1], EUR (n=9), and MIX [n=6] populations. The CSA participant was homozygous for V456A, but various CF-causing variant combinations were found in the EUR and MIX populations. F508del was the most common CF-causing INDEL detected across the populations. Interestingly, the two individuals homozygous for F508del were also diagnosed as having CF in UKBB. Many of the participants with two CF-causing *CFTR* variants had combinations associated with pancreatic sufficiency (96%). The three variant combinations associated with pancreatic insufficiency were R851X/F508del and F508del/F508del [n=2] (Table 3). We found 155 participants that harbored a CF-causing variant and a VCC (Table 4). These participants were found only in the EUR [n=145] and MIX [n=10] populations. R117H was the most reported VCC [49/154] variant in this subset of participants, and its combination with F508del was the most common found in the EUR [38/145] and MIX [5/10] populations. L967S also featured as a common VCC variant. Its combination with F508del was exclusively reported in the EUR population and dominated as the second most common combination (Table 4). Since high impact variants are potentially functionally relevant, we assessed their distribution across the ancestries. Among the 200 high impact variants detected in the participants, the highest number was found in EUR population [150/200] while the least number of these variants [4/200] were reported in both the AMR and EAS populations [Table S7]. We also found a high number of participants heterozygous for a CF-causing and high impact variant [Table S9]. These findings warranted us to interrogate the association of these variant with participants health outcomes.

**Table 3.**
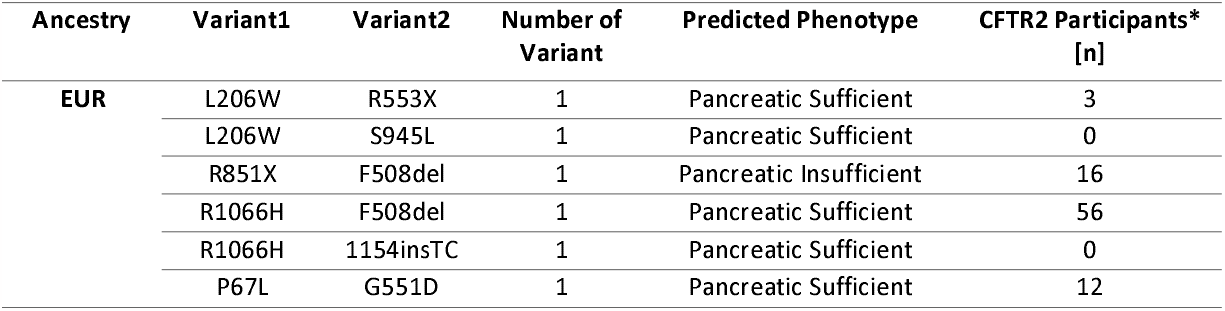
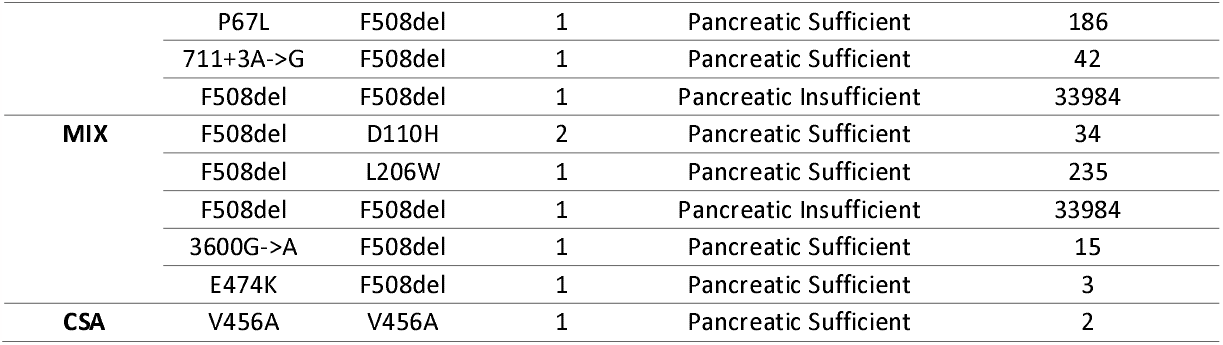
Participants with two CF-causing variants

**Table 4.** UKBB participants with a CF-causing and variable clinical consequence (VCC) variant

| Ancestry | Variant1 | Variant2 | Count |
| --- | --- | --- | --- |
| **EUR** | R117H | F508del | 38 |
|  | L967S | F508del | 30 |
|  | D443Y | F508del | 13 |
|  | P750L | F508del | 12 |
|  | 621+3A->G | F508del | 5 |
|  | R334Q | F508del | 4 |
|  | P750L | G551D | 3 |
|  | R258G | F508del | 3 |
|  | D1152H | F508del | 2 |
|  | F1052V | F508del | 2 |
|  | L967S | 1717-1G->A | 2 |
|  | L967S | G551D | 2 |
|  | L967S | W1282X | 2 |
|  | Q1291H | F508del | 2 |
|  | R1070W | F508del | 2 |
|  | D1270N | F508del | 1 |
|  | D443Y | 1898+1G->A | 1 |
|  | D443Y | N1303K | 1 |
|  | D443Y | Q493X | 1 |
|  | D443Y | R553X | 1 |
|  | F575Y | 1154insTC | 1 |
|  | L967S | 1138insG | 1 |
|  | L967S | 1898+1G->A | 1 |
|  | L967S | 3659delC | 1 |
|  | M265R | F508del | 1 |
|  | P750L | 1461ins4 | 1 |
|  | P750L | R560T | 1 |
|  | P750L | W846X | 1 |
|  | R1070Q | F508del | 1 |
|  | R1070W | G542X | 1 |
|  | R117G | F508del | 1 |
|  | R117H | 1898+1G->A | 1 |
|  | R117H | 2184delA | 1 |
|  | R117H | 3272-26A->G | 1 |
|  | R117H | I507del | 1 |
|  | R117H | R347H | 1 |
|  | R117H | R347P | 1 |
|  | R352W | N1303K | 1 |
| MIX | R117H | F508del | 5 |
|  | 621+3A->G | F508del | 1 |
|  | D443Y | F508del | 1 |
|  | D1152H | 1898+1G->A | 1 |
|  | F1052V | F508del | 1 |
|  | Q1476X | N1303K | 1 |

### Classical CF phenotypes reported in participants with two CF-causing variants and VCCs

Phenotype analysis performed using hospital in-patient records indicated that most of the participants with at least two CF-causing variants presented classical CF phenotypes. Eight (International Classification of Diseases 10^th^ Revision (ICD-10) codes were significantly [FDR<0.05] enriched in participants with two CF-causing variants (Fig. 5A). Specifically, ICD codes directly associated with CF [E840, 848, and 849] and colonization with *Pseudomonas aeruginosa* [B965] were among the top significantly enriched terms. Interestingly, ICD Code U837 which corresponds to Resistance to Multiple Antibiotics also featured among the top 10 enriched phenotypes. Although bronchiectasis, a classical consequence of dysfunctional CFTR, was listed among the top, it did not meet our significance threshold when corrected for multiple testing (Fig. 4). With VCC variants resulting in variable clinical outcomes when combined with another CF-causing variant but not often diagnosed as CF, we assessed the phenotypes of participants that fall within this category. Our analysis indicated that six ICD codes were significantly [FDR<0.05] associated with CF [E840 and 848]; of these, pulmonary phenotypes [J47, B441, J998, and J440] were significantly enriched in these participants (Fig. 5B). In an attempt to characterize the clinical relevance of high impact variants, we then explored the phenotypes of participants with one CF-causing variant and a high impact variant not known to cause CF (n=272). Surprisingly, the only significant association [FDR<0.05] returned was Melanoma in situ of the trunk which is not a classical CF phenotype. Aspergillosis was featured among the top 10 most enriched phenotypes, but this association failed to meet our significance threshold (Table S9). Further analysis deployed to compare differences of spirometry measures (FEV1 and FVC) between participant with two CF-causing variants and the general population returned no significant results.

**Fig. 5:**
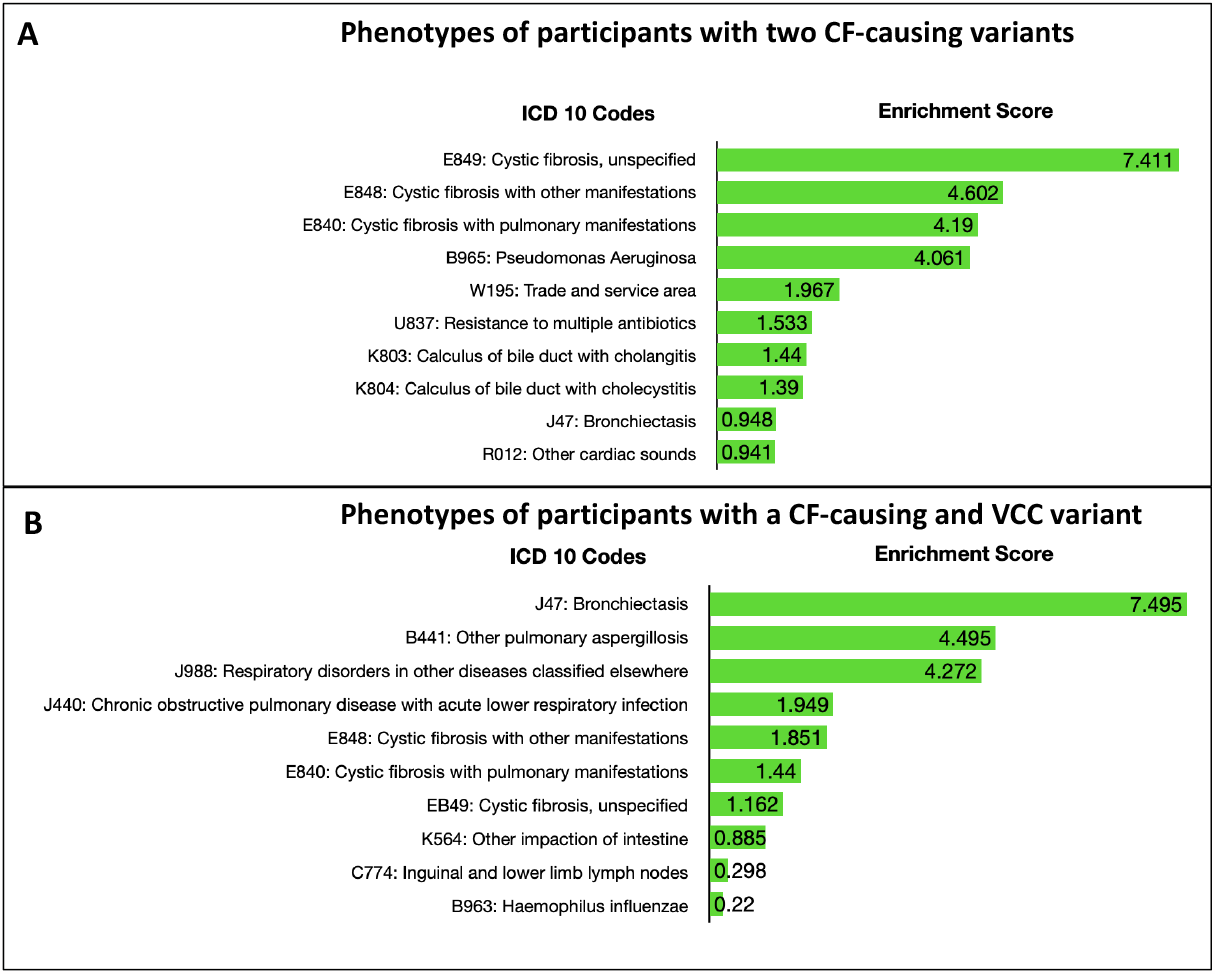
Classical CF phenotypes enriched in participants with >1 clinically relevant *CFTR* variant. (A)Enriched phenotypes observed in patients with two CF-causing variants **B)** Enriched phenotypes observed in patients with one CF-causing and a high impact variant. The top 10 enriched phenotypes are represented in bar graphs. The negative logarithm of the Bonferroni adjusted p-value was used to deduce the enrichment scores shown in the bars. Enrichment score >1.3 corresponds to a significance threshold of *p*<0.05.

### Chronological age of CF-causing variants

GEVA reported the estimated age of over 4,000 variants within the *CFTR* gene locus (Table S10). Only four of the dated variants were CF-causing [rs139729994 (3600G>A), rs193922500 (V456A), rs76713772 (1717-1G>A), rs77646904 (V520F)] (Fig. S1). The youngest variant among these, 1717-1G->A, appears to have originated ∼425 generations ago (∼10,600 years) while the earliest variant, V456A, originated about ∼215 generations ago (∼5,300 years). Interestingly, V520F and 3600G>A which appeared ∼338 and 270 generations ago featured among the most common CF-causing variants reported in EAS population (Fig. 3). When the allele frequencies of the CF-causing variants were compared with variants of similar ages within the respective populations in AVA, there were no significant differences. Thus, an indication that the variants were not under any selective pressure at the time of origination.

## Discussion

Molecular diagnosis of CF is confounded by sparse knowledge about the prevalence of pathogenic and non-pathogenic *CFTR* variants across ancestries, including those that are potentially ancestry specific. In this study, we interrogated whole-exome sequence datasets generated from over 450, 000 UKBB participants to characterize the distribution of *CFTR* variants across six ancestries [AFR, CSA, EAS, EUR, AMR, and MID], including an uncharacterized group (MIX). We report, for the first time, the detection of over 4000 *CFTR* variants across the ancestries. Among the detected ancestry-specific variants were several variants of clinical and functional relevance. Phenotypic characterization of participants harboring multiple combinations of these variants reported indications associated with CF and its classical pulmonary phenotypes.

Although previous efforts driven by the CFTR1/2 team have led to the identification of just over 2000 *CFTR* variants within the CF population^10^, our report of over 4000 *CFTR* variants is a small fraction of potential variants that could exist within the CFTR gene. *In silico* analyses of external variant browsers such as BRAVO^17^, which is based on ∼132k WGS generated by the TOPMed consortium, and All of Us Research Hub^25^, which is based on ∼98k WGS and ∼165k genotyping arrays reports a total of 56,448 and 39,797 *CFTR* variants, respectively. The higher number of *CFTR* variants captured in these databases are perhaps due to WGS having the capability of capturing the entire length of the *CFTR* gene while WES captures mostly exonic regions. Overall, the vast majority of the *CFTR* variants found in UKBB [52%], All of Us Research Hub [89%], and BRAVO [96%], were intronic variants. Overall, c.1408G>A (V470M [rs213950]) was the most common allele (Table S1). Although in GRCH37/38 the reference allele is a G, the variation associated with increased risk of chronic pancreatitis indicate at this position is c.1408A>G (M470V)^26^. Regardless, a combination of this variant with another CF-causing does not cause CF (https://cftr2.org). CF genetic diagnosis is largely based on characterized *CFTR* variants, most of which are reported in Europeans^4,10^. Although the UKBB is dominated by people of European ancestry, which could potentially influence estimates of genetic variations^27^, CF is one of the most common life-threatening genetic disease reported in this population28. In line, with previous reports we also found the highest number of *CFTR* variants in EUR population. Thus, the mutation rate of the *CFTR* gene is likely higher in the EUR than other populations. The aspect of precision medicine for CFTR modulators is bound to make higher impact in this CF population with such variable numbers of mutations.

Many clinically (CF-causing and VCC) and functionally (High-Impact) relevant variants were identified following cross-ancestry annotation of all detected variants. As their global distribution and frequencies are poorly understood, we characterized the most common and unique CF-causing variants across populations. Indeed, like previous reports^13,29,30^, F508del was the most common CF-causing variant reported globally. While F508del represented ∼90% of the CF-causing variants found in EUR, our findings indicate its prevalence is much lower in other populations (Fig. 3). Thus, F508del may be the major cause of CF in EUR population, but not for people of all ancestries. Lower frequencies of F508del in non-European populations have been reported in several studies, and the spectrum of variants causing CF in such populations varies^31,32^. For example, G970D has been reported as the most common cause of CF in the Chinese population^12^. This variant, along with V520F and 3600G>A, featured among the top three common variants we found in the EAS group and were more common than F508del (Fig.3). Although we found V520F to be the most common in EAS, it is conceivable G970D is more common to China than V520F since the EAS group examined in this study expands beyond the Chinese population. Like other non-EUR populations, CF is less frequent or likely underreported in Africans10. Although F508del was the most common CF-causing variant found in the AFR group, 3120+1G>A which is predominantly found in Africans^10^, was also found to be common to the AFR group (Fig. 3). Also, in South Asians, CF is thought to be less frequent. Although F508del represents about 40-50% of cases, V456A the second most common CF-causing variant we found in CSA (Fig.3), is a well-characterized cause of CF in South Asians^33,34^. Some other common variants found in non-EUR populations, such as AFR (3791delC), CSA (1525-1G>A), and EAS, were ancestry-specific. By comparing our findings with BRAVO reports^17^, we found these variants were also rare and exclusively possessed by similar populations. Unfortunately, we found most of the common CF-causing variants found in non-European populations are not included in the American College of Medical Genetics list of 23 variants recommended for CF carrier screening^35^.

Numerous hypotheses have been put forward to explain the persistence of CF-causing variants across populations, despite their detrimental impact in life outcomes. A leading hypothesis postulates the heterozygous advantage of CF-causing variants against infectious diseases, such as cholera, typhoid fever, or tuberculosis36. For example, the most common CF-causing variant, rs113993960 (F508del), has been estimated to have arisen approximately 600 generations ago^37,38^ and its rapid increase in Europeans (MAF = 1.6%) was linked to the tuberculosis pandemic of the seventeenth century, against which it is thought to increase resistance in mutation carriers^36^. Among other variants with estimated age data in AVA, we did not detect any variant that has rapidly increased in allele frequency as the F508del variant in Europeans. Although, V520F and 3600G>A are more common CF-causing variants in EAS population than F508del (Fig. 3), their allele frequencies were not significantly different from other variants of similar ages in EAS population. Thus, F508del is likely a unique variant that confers selective pressure, which has kept its frequency relatively high.

Beyond CF-causing variants, we also found several VCCs, and High Impact variants not known to cause CF but predicted to severely impact protein function. R117H the common VCC, which results in reduced single-channel activity and open probability^39^, was the most prevalent VCC detected across the ancestries (Table S5). Clinical manifestations of patients with R117H are heterogenous and largely influenced by its combination with another CF-causing variant or other common variants of poly-T tract (5T, 7T, and 9T). R117H-T5 is reported to potentially result in less functional CFTR and pancreatic sufficiency when patients are homozygous for this combination or when found in compound heterozygosity with a CF-causing variant40. Most of the predicted High Impact variants found in our study were already known to cause CF (deduced with the CFTR2 annotation). After excluding these variants, we still found some uncharacterized High Impact variants. Taken together, these findings indicated a handful of UKBB participants harbored clinically and functionally relevant *CFTR* variants.

Since CF is an autosomal recessive disease, we characterized UKBB participants with more than one clinical or functional relevant variant. Interestingly, we found sixteen UKBB participants with at least two CF-causing variants. Collectively, these participants reported ICD codes associated with classical CF phenotypes [Fig. 5A], despite the fact only two participants (F508del homozygotes) were diagnosed as CF in UKBB electronic health records. Undiagnosed cases of CF, especially in patients with milder forms of the disease caused by rare pathogenic genotypes, is not uncommon^41^. Most of the variant combinations reported in our findings likely result in pancreatic sufficiency, except for one participant harboring the F508del/R851X genotype that results in pancreatic insufficiency. Although the F508del/R851X participant was not diagnosed as CF in UKBB, the retrieved ICD10 code indicated the participant suffered majorly from Chest Pain [R074]. This variant combination, which was detected in an EUR participant, is less common in CF, with a frequency of only ∼0.0002 recorded in CFTR2 database [https://cftr2.org]. Another rarer genotype with ∼0.00002 frequency in CFTR2 which we found was V456A/V456A. This variant is known to cause CF in South Asians^33^ and likewise was detected in a CSA participant. Some of the detected pathogenic genotypes, such as L206W/S945L and R1066H/1154insTC, have not yet been recorded in CFTR2. Taken together, a possible explanation for the undiagnosed cases captured in our study may be due to the rareness of the variants and milder presentation of the disease.

We recorded about 10-fold more participants harboring a CF-causing variant and a VCC, than those with two CF-causing variants (154 vs 16, respectively). VCCs contribute majorly to the complexities associated with genetic diagnosis of CF, because clinical outcomes vary widely across patient populations^42^. For this sub-group of participants, we also found significant enrichment of ICD codes associated with classical CF (ICD Codes: E840 and E848) and pulmonary phenotypes, including bronchiectasis, Aspergillosis, and COPD with acute lower respiratory infections. Participants with these genotypes were reported only in EUR and MIX populations, and the most dominant combination was F508del/R117H. This variant combination on its own does not cause CF but when in *cis* with 5T ploy-Tract variant, then CF diagnosis is likely^40^. About 1,310 patients have the F508del/R117H variant combination in CFTR2 and 22% of these are pancreatic insufficient. Meanwhile, 80 patients have the F508del/R117H;5T combination and 33% of them are pancreatic insufficient [https://cftr2.org]. Although these participants are not diagnosed as CF, their inclusion as patients in CFTR2 suggest they suffer the burden of dysfunctional CFTR. The F508del/L967S genotype, which was the second most common found in this subgroup (CF-causing+VCC), was rare in CFTR2. Only seven patients had this combo and clinical outcome, though variable, is largely expected to be pancreatic sufficient. The identification of several UKBB participants harboring such variant combinations and showing CF-like symptoms, is therefore an indication that a number of undiagnosed or unreported CF cases may benefit from modulator therapies. Additionally, we found no differences in pulmonary measures (FEV and FVC) between participants with two CF-causing variants and the general population. Given that the UKBB recruited individuals that were over 40 years old, the individuals with multiple clinically relevant *CFTR* genotypes likely have a milder form of the disease with a different disease etiology that nevertheless could be potentially either treated or prevented by appropriate CFTR-based interventions.

A major drawback to existing knowledge about pathogenic *CFTR* variants is that most studies have been conducted in European populations. However, CF is also prevalent in people from other ancestries, who may harbor uncharacterized unique variants4. Even in individuals of European ancestry, where most of the studies have been conducted, there are several *CFTR* variants with unknown clinical significance. In attempt to decipher the clinical relevance of uncharacterized high impact *CFTR* variants detected across the UKBB population, we performed a phenotype analysis of participants with a CF-causing variant and a non-CF-causing (uncharacterized) high impact variant but found no interesting association. Databases such as the UKBB offer an unprecedented opportunity to correlate uncharacterized genotype and phenotype information at a population scale. Although we utilized the UKBB resource to characterize the ancestral diversity of *CFTR* variants, most of the participants are of European ancestry. Recent genome sequencing efforts that included more diverse populations, such as the NIH All of Us and ToPMed programs, report even more *CFTR* variants in comparison to UKBB. However, limited access to phenotype information at an individual level makes it challenging to unravel the clinical relevance of the detected *CFTR* variants.

Here, we aimed to characterize the natural variation that exists within the *CFTR* gene that is not limited to persons with CF. The large majority (>95%) of variants surveyed have allele frequencies less than 1% and ∼9% of all variations are indels. While sequencing technology has improved in accuracy and quality control filters have removed many of the problematic variants, identification, and interpretation of novel rare variants (specifically singletons and doubletons) would warrant more precise methods (such as targeted sequencing) for confirmation. Another limitation of our study is the lack of precise CF-related phenotypes, such as FEV1% predicted and sweat chloride levels in the electronic health records extracted from UKBB. Although we captured some spirometry measures, we lacked information relevant to deduce FEV1% scores and there were no records of sweat chloride levels for the participants.

In summary, we have leveraged the UKBB resource to comprehensively characterize the broad spectrum of *CFTR* variants across ancestries. Until now, most efforts aimed at identifying and characterizing *CFTR* variants have focused more on surveying CF populations in a relatively smaller sample size. For the first time, we report the detection of over 4000 *CFTR* variants, which nearly doubles the number of variants reported by CFTR1/2. However, the higher number of variants reported by the NIH All of Us and ToPMed diverse sequencing programs suggests the 4000 *CFTR* variants represents only a small fraction of variants that exist in the *CFTR* gene. The identification of several ancestry-specific variants, including uncharacterized functionally relevant variants, warrants the need for CF screening panels to consider ethnic specificities. The presentation of classical CF phenotypes seen in some non-CF diagnosed participants, with CF-causing and VCC variants, indicates they may benefit from current *CFTR* therapies. As more diverse *CFTR* sequences are becoming increasingly available in genomic databases, future studies are encouraged to leverage such resources to characterize the clinical relevance of high impact variants not previously known to cause CF.

## Data Availability

Access to UK biobank resource is available by application (http://www.ukbiobank.ac.uk/). The UK biobank exome sequences analyzed in this study were obtained under application 26401.

## Supplementary information

Supplementary Tables 1-7: **Table S1**. Detected *CFTR* variants across UKBB populations; **Table S2**. Unique CF-causing variants; **Table S3**. CF-causing *CFTR* variants; **Table S4**. Variable clinical consequence variants detected across UKBB ancestries; **Table S5**. Unique CF-causing variants; **Table S6**. Common varying clinical consequences (VCC) variants; **Table S7**. *CFTR* variants annotated as having High Impact; **Table S8**. Uncharacterized high impact variants; **Table S9**. High impact and CF-Causing variants. **Table S10**. Dated variants within the CFTR gene locus

Supplementary Figures: **Figure S1**. Venn diagram showing genealogical estimation of variant age (GEVA) analysis of CF-causing variants

## Ethics declarations

Competing interests: The authors declare no competing interest. The design, study conduct, and financial support for this research were provided by AbbVie. AbbVie participated in the interpretation of data, review, and approval of the publication. JEI, ML, BRG, SRP, FR, PK, RT, PD, AS, JFW, and AV are employees of AbbVie. HL is an employee of University of Wisconsin School of Medicine and Public Health and provided her expertise on CF genetics and translational research (no funding to disclose).

## Acknowledgement

We would like to acknowledge and extend our gratitude to the AbbVie GRC Internal Reviewers; Anneke Den Hollander and Nizar Smaoui.

