## Supplementary material for "Diversity of *CFTR* variants across ancestries characterized using 454,727 UK biobank whole exome sequences": Figure S1

**Fig. S1**. Venn diagram showing genealogical estimation of variant age (GEVA) analysis of CF-causing variants.

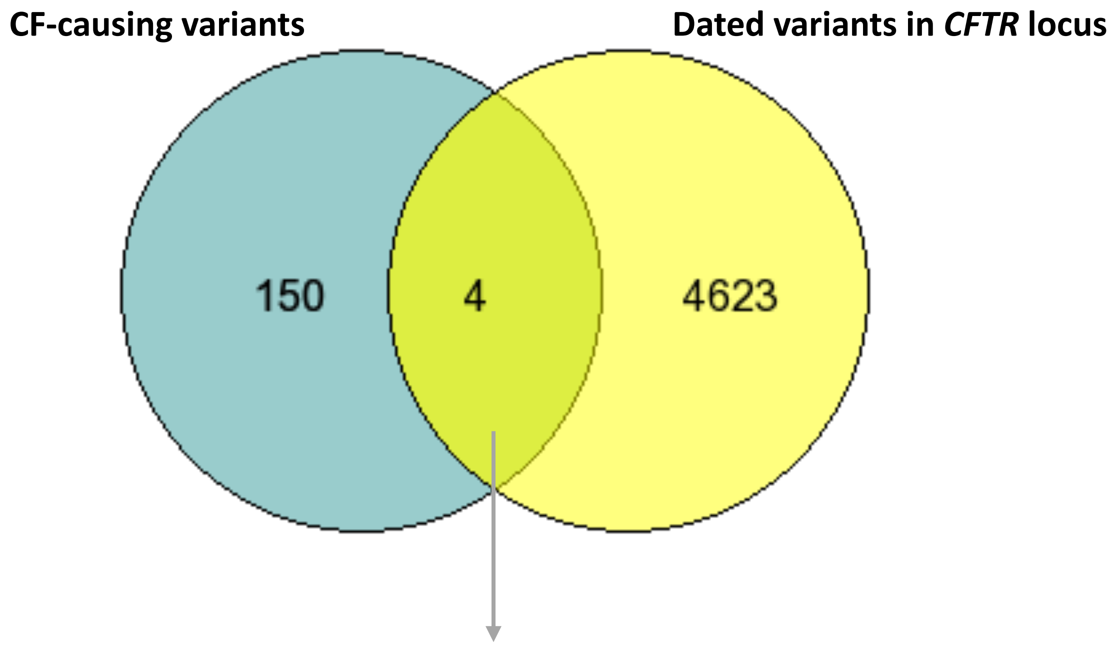


| rsID | Variant Name | Legacy Name | Allele Age (years) | Time (Generations) |
|---|---|---|---|---|
| rs139729994 | c.3468G>A | 3600G>A | 6700 | 270 |
| rs193922500 | c.1367T>C | V456A | 5300 | 215 |
| rs76713772 | c.1585-1G>A | 1717-1G->A | 10,600 | 425 |
| rs77646904 | c.1558G>T | V520F | 8500 | 338 |

Age estimates of four CF-causing variants dated in GEVA are shown. A total 4/4627 *CFTR* variants dated in GEVA are CF-causing. 1717-1G>A was the oldest among all four dated CF-causing variants while V456A was the youngest.
